# Essential epidemiological parameters of COVID-19 for clinical and mathematical modeling purposes: a rapid review and meta-analysis

**DOI:** 10.1101/2020.06.17.20133587

**Authors:** Eva S. Fonfría, María Isabel Vigo, David García-García, Zaida Herrador, Miriam Navarro, Cesar Bordehore

**Author notes:** Corresponding author: Dr. Cesar Bordehore, Department of Ecology, University of Alicante, Campus San Vicente del Raspeig, 03690, Spain. Alternate corresponding author: Dr. Eva Fonfría, “Ramon Margalef” Environmental Research Institute (IMEM), University of Alicante, Campus San Vicente del Raspeig, 03690, Spain.

## Abstract

We present a literature review and meta-analysis of relevant epidemiological parameters (24 for adults, 7 for children) of COVID-19. Standardization of these parameters is key to performing valid clinical and mathematical modeling, as well as forecasts, helping us to improve our understanding about the characteristics and impact of the pandemic.

## Text

Without any doubt, the coronavirus disease 2019 (COVID-19) outbreak, caused by the novel coronavirus SARS-CoV-2, is the most significant global public health threat in recent decades. It was declared as “Public Health Emergency of International Concern” and “pandemic” by the World Health Organization (WHO) in 30 January and 11 March 2020, respectively. Since it was first detected December 2019 in Wuhan, China, it has spread over 212 countries and territories, with almost 6,800,000 confirmed cases and 400,000 deaths worldwide as of 7 June 2020 [1].

In the absence of effective treatments or preventative therapeutic strategies for COVID-19 [2], epidemiological models have become key tools for policymakers and stakeholders to estimate the probable time course of the pandemic and to evaluate the effectiveness of the measures adopted to manage it [3–5]. Since the beginning of the pandemic, thousands of preprints and papers have been published about COVID-19. However, sample sizes were usually too small to be able to generalize the values and to enable better models and evidence-based clinical decisions. It is desirable that those parameters be defined and determined as accurately as possible, taking into account the inter-study variability with the appropriate mathematical methodology. Besides, most studies so far examine only a handful of variables, making data mining particularly onerous and tedious. The aim of this study is to identify and summarize the key epidemiological parameters used so far for mathematical modeling and clinical purposes.

We searched PubMed, MedRxiv and BioRxiv for articles and preprints published in English between 1 January and 15 April 2020 with the keywords “COVID-19”, “SARS-CoV-2”, “2019-nCoV” or “novel coronavirus” as well as terms to obtain data about the following parameters (definitions and reclassifications are described in the Supplementary Material): *percentage of presymptomatic transmissions, presymptomatic transmission period, percentage of asymptomatic patients, serial interval, incubation period, onset of symptoms/illness onset to diagnosis, onset of symptoms/illness onset to hospital admission, hospital stay length, hospital admission to death, hospital admission to discharge, onset of symptoms/illness onset to death, onset of symptoms/illness onset to discharge/recovery, percentage of deaths, percentage of discharged, percentage in hospital, percentage of Intensive Care Unit (ICU) admissions, onset of symptoms/illness onset to ICU admission, ICU stay length, percentage of deaths from ICU, percentage of discharged from ICU, percentage in hospital from ICU, and percentage transferred from ICU to general hospital wards*.

A preliminary screening by title and abstract was conducted to identify potentially relevant studies. Subsequently, the full texts of those studies were evaluated and their reference lists examined for additional records (see Supplementary Figure 1 for the flow diagram). Relevant epidemiological parameters, such as sample size, mean, standard deviation (SD), confidence interval (CI), median, interquartile range (IQR) and the fitted distribution used in its estimation, when applicable, as well as sociodemographic information (i.e. patient’s age, gender and location) were extracted. Reviews and non-original research papers were excluded. In case of data overlap (e.g. several studies reported data about “asymptomatic patients” among passengers of the Diamond Princess cruise ship), the article with the largest sample size was chosen. Finally, because COVID-19 does not seem to affect children and teens in the same way as adults [6], we decided to analyze the obtained data for pediatric patients separately. Initial screening was performed by ESF, followed by an ultimate extraction of data assessed independently by ESF, DGG and MIV. When discrepancies were detected, MIV made the final inclusion decision.

To integrate all the reviewed information in one overall estimate that could be introduced in mathematical models, we performed different meta-analysis (see Supplementary Material for detailed information). For those parameters including mean and SD, or median and IQR, we carried out a meta-analysis of single means; when mean and SD were not reported, they were estimated from median and IQR [7,8]. For those parameters presented as percentages, a meta-analysis of proportions was implemented. To take into account the variability between and within studies, *random-effects* models were fitted with the Restricted Maximum Likelihood Method (REML). To meet the normality assumption underlying the meta-analysis, the natural logarithm transformation was applied. The null hypothesis of no variance among studies (τ^2^ =0) was tested using the Q-statistic, and the degree of heterogeneity was quantified by the I^2^ index [9]. Outliers and influencers diagnoses were also performed [10]. Data were analyzed using the statistical software R version 3.6.2 and the “meta”, “metaphor” and “dmetar” packages.

Between 1 January and 15 April 2020, 6,969 scientific articles related to COVID-19 were published in Pubmed or posted in MedRxiv and BioRxiv preprint servers. After filtering using the aforementioned keywords and terms and performing the initial screening of titles and abstracts, a total of 343 papers were reviewed in full text. From them, 126 contained extractable data (see Supplementary Table 1), 118 were considered initially in our meta-analysis for one or more parameters and after removing outliers/influencers, 102 were included in the final analysis. Table 1 shows the pooled mean or percentage and its associated 95% CI provided by the meta-analysis for parameters with three or more studies, i.e., 18 parameters (3 of them disaggregated in survivor/non-survivor) for the general population and 7 for children. For each parameter, forest plots to visualize results when all the studies were included in the meta-analysis and with outliers and/or influencers removed are provided in the Supplementary Material (Supplementary Figures 2-32). For most parameters, the heterogeneity was moderate to high.

**Table 1.** Meta-analysis results.

| Parameter | Mean | 95% CI | Studies included* | N (patients) |
| --- | --- | --- | --- | --- |
| **General population** |  |  |  |  |
| Serial interval (days) | 5.06 | 4.50-5.70 | 11 | 852 |
| Incubation period (days) | 5.79 | 5.48-6.11 | 21 | 1792 |
| Onset of symptoms to diagnosis (days) | 5.82 | 4.74-7.15 | 7 | 814 |
| Onset of symptoms to hospital admission (all) (days) | 6.49 | 5.94-7.10 | 14 | 1544 |
| Onset of symptoms to hospital admission (survivor) (days) | 6.60 | 4.23-10.29 | 3 | 314 |
| Onset of symptoms to hospital admission (non-survivor) (days) | 10.18 | 9.65-10.74 | 6 | 569 |
| Onset of symptoms to hospital ICU admission (all) (days) | 9.69 | 9.14-10.26 | 5 | 174 |
| Hospital stay length (all) (days) | 14.21 | 13.38-15.10 | 7 | 991 |
| Hospital stay length (non-survivor) (days) | 9.16 | 8.29-10.13 | 9 | 453 |
| Hospital stay length (survivor) (days) | 15.21 | 14.37-16.10 | 12 | 737 |
| ICU stay length (all) ** (days) | 8.72 | 7.57-10.05 | 3 | 414 |
| ICU stay length (non-survivor) (days) | 8.16 | 7.26-9.17 | 5 | 371 |
| ICU stay length (survivor) (days) | 10.91 | 9.20-12.94 | 4 | 87 |
| Onset of symptoms to death (days) | 16.71 | 15.37-18.17 | 11 | 601 |
| Onset of symptom to discharge ** (days) | 21.83 | 19.20-24.85 | 4 | 522 |
| Presymptomatic transmission (%) | 39.04 | 18.38-64.56 | 4 | 713 |
| Asymptomatic patients (%) | 8.99 | 5.58-14.18 | 17 | 2304 |
| ICU admissions vs all COVID-19 hospital admissions (%) | 18.16 | 15.74-20.86 | 12 | 5877 |
| Deaths vs all hospitalized patients with COVID-19 (%) | 9.16 | 6.91-12.03 | 14 | 2608 |
| Deaths from ICU (%) | 29.17 | 14.69-49.62 | 8 | 555 |
| Discharged vs all hospitalized patients with COVID-19 (%) | 42.79 | 36.76-49.05 | 15 | 4064 |
| Discharged from ICU (%) | 35.31 | 21.56-52.00 | 9 | 565 |
| In hospital (%) | 53.92 | 46.81-60.86 | 16 | 4181 |
| In hospital (ICU) (%) | 20.14 | 12.29-31.21 | 8 | 238 |
| **Children (<18 years)** |  |  |  |  |
| Incubation period ** (days) | 6.69 | 5.49-8.15 | 4 | 25 |
| Onset of symptoms to diagnosis ** (days) | 3.27 | 1.35-7.92 | 3 | 744 |
| Hospital stay length ** (days) | 12.77 | 8.51-19.17 | 3 | 52 |
| Asymptomatic patients ** (%) | 16.66 | 11.52-23.48 | 4 | 945 |
| ICU admissions vs all COVID-19 hospital admissions ** (%) | 5.06 | 1.64-14.60 | 4 | 211 |
| Discharged vs all hospitalized patients with COVID-19 ** (%) | 19.39 | 3.39-62.23 | 3 | 205 |
| In hospital ** (%) | 80.27 | 37.87-96.45 | 3 | 205 |

Abbreviations: CI, Confidence Interval; ICU, Intensive Care Unit.
\* References are compiled in the Supplementary Appendix (Table S2).
\*\* Results are based in less than 5 studies and they have to be interpreted cautiously. Outliers and influencers analysis is not included because it is useless.

Compared to other available reviews on COVID-19 [11,12], our study summarized a larger number of epidemiological parameters and covered a longer period of time, substantially increasing the number of studies considered per parameter. This study also has several limitations. First, only papers in the English language were evaluated. Although many papers have been published in Chinese and a few in other languages, papers in English provided a large enough sample. Second, we performed the search strategy only in three libraries and up to 15 April 2020. We realize our work might not include all the published data with the selected parameters; however, the number of papers about COVID-19 posted daily is so large that any attempt to perform an in-depth systematic review would be rapidly outdated. Finally, we included preprints, which, although they had not yet been peer-reviewed and their results should be interpreted cautiously, contain valuable information.

In summary, we present a review of the most relevant epidemiological parameters of COVID-19 so far. Our results will promote reliable and more accurate forecasts using mathematical modeling of the epidemic itself and also of the material and human resources needed. These data may also be of interest to clinicians, e.g. for their daily work, interpretation of clinical evolution, or clinical trials design. We expect it to be useful to other modelers, managers, and national and regional policy makers when deciding the appropriate mitigation strategies for the ongoing global pandemic.

## Data Availability

All data are offered in the paper and references in it.

## Funding

This work was supported by the University of Alicante [COVID-19 2020-41.30.6P.0016 to CB] and the Ajuntament de Dénia through the Montó-Dénia Research Station Agreement [2020-41.30.6O.00.01 to CB].

## Acknowledgements

We acknowledge Parques Nacionales (Ministerio para la Transición Ecológica y Reto Demográfico, Spain) and Generalitat Valenciana (Regional Government of Valencia, Spain) for the support of the Montgó-Dénia Research Station. Editing was provided by Sea Pen Scientific Writing.

## Notes

### Competing Interest Statement

The authors have declared no competing interest.

### Funding Statement

This work was supported by the University of Alicante [COVID-19 2020-41.30.6P.0016 to CB] and the Ajuntament de Denia through the Montg-Denia Research Station Agreement [2020-41.30.6O.00.01 to CB].

### Author Declarations

University of Alicante

